# Associations and Interaction Effects of Socioeconomic, Lifestyle, and Genetic Factors on Intrinsic Capacity

**DOI:** 10.1101/2025.10.05.25337351

**Authors:** Melkamu Bedimo Beyene, Renuka Visvanathan, Robel Alemu, Olga Theou, Beben Benyamin, Matteo Cesari, John Beard, Azmeraw T. Amare

## Abstract

**Background:** Intrinsic capacity (IC) is a composite measure, computed from five domains: cognition, psychological well-being, locomotion, vitality, and sensory. IC reflects the overall physiological reserve and functional capacity of an individual, making it a key indicator of healthy ageing. The substantial interindividual variability in IC is likely influenced by genetic (polygenic) as well as socioeconomic status and lifestyle factors. However, the interaction effect of these factors is yet to be explored.

**Objective:** This study examined (1) associations of IC with socio-economic and lifestyle factors and the polygenic scores for IC (PGS-IC) when stratified by age, and (3) the interaction effects of the PGS-IC and socio-economic or lifestyle factors on IC.

**Methods:** Our study included 13,112 participants from the Canadian Longitudinal Study on Aging (CLSA) comprehensive cohort with complete IC variables and genetic data. Composite lifestyle scores, including the Physical Activity Scale for the Elderly (PASE), Prospective Urban Rural Epidemiological study (PURE) diet, and Mediterranean diet scores, were generated following established guidelines. Associations of IC with the socioeconomic and lifestyle factors were assessed using linear regression models adjusted for age and sex. The IC scores and the PGS-IC were developed in CLSA in our previous work, and this study tested age-stratified associations of PGS-IC with IC, and interaction effects of the PGS-IC and socioeconomic or lifestyle factors on IC using linear regression models adjusted for age, sex, and the top five genetic principal components. Statistical significance was defined as a false discovery rate (FDR) adjusted *P* < 0.05.

**Results:** The mean age was 61 (standard deviation 9.6) years, and 50.8% were females. Higher IC was associated with higher education (B = 0.255, 95% CI: 0.180, 0.329), higher income (B = 0.392, CI: 0.322, 0.461), physical activity (PASE score: B = 0.001, CI: 0.0004, 0.001), and healthier diets (PURE diet score: B = 0.024, CI: 0.021, 0.027; Mediterranean diet score: B = 0.018, CI: 0.016, 0.021). IC was lower in previous (B = -0.093, CI: -0.121, -0.064) and current smokers (B = -0.407, CI: -0.459, -0.355) compared to never smokers. Likewise, short (<7h: B = -0.133, CI: -0.161, -0.105) and long (>9h: B = -0.258, CI: -0.392, -0.124) sleep durations were negatively associated with IC compared to those who had optimal sleep. The PGS-IC was positively associated with IC, particularly in younger adults. Significant interaction effects were observed with Mediterranean diet (B = -0.003, CI:-0.006 -, -0.0002) in whole sample, education in younger adults (B = -0.109, CI: -0.211 -, -0.007), and sleep (younger adults: long sleep, B = 0.198, CI:0.023, 0.373; older adults: short sleep, B = -0.095, CI: -0.153 -, -0.036).

**Conclusion:** Novel findings confirming the interaction effects of PGS-IC with socioeconomic and lifestyle factors suggest that there is a complex interplay between genetics and the environment in shaping IC and healthy ageing.

## 1. Introduction

With population ageing observed globally at an unprecedented rate, maintaining health and functionality in older age is a public health priority [1]. Traditional approaches to ageing have predominantly focused on disease management and morbidity reduction. However, there is a growing consensus that a broader preventative approach across the lifespan is needed - one that maximizes functional ability in older age, enabling older people to be functionally able, continuing to do what they value as they age [1–3]. The World Health Organization (WHO), within its healthy ageing framework, outlined that functional ability is influenced by IC, the environment (e.g., socio-economic and lifestyle factors), and the interaction of these two factors [1, 4]. IC is operationalised through five key domains - locomotion, vitality, cognition, psychological well-being, and sensory function [4–7].

There is growing evidence that IC itself is influenced by a dynamic interplay between genetic predisposition and external factors, including socioeconomic, lifestyle, and behavioural influences [5, 8, 9]. Our scoping review [10] identified a wide range of socioeconomic and lifestyle factors associated with IC, such as educational, social, and economic status, engagement in social activities, and other lifestyle factors such as diet, exercise, physical activity, smoking, and sleep. Our research program using CLSA and UK biobank data suggested that a quarter of the interindividual variability in IC is attributable to genetic variation, and that the PGS-IC in CLSA was associated with the IC scores [8]. Understanding the consistency of PGS-IC across age groups and how they interact with socioeconomic and lifestyle factors to influence IC is an important next step in our program of research, as it will help identify potential targets for intervention, modifiable factors that could help preserve IC, and promote healthy ageing.

The primary aim of this study was to leverage data from the CLSA to examine the association between IC and socio-economic and lifestyle factors, and the PGS-IC when stratified by age, and explore the interaction effects of PGS-IC and the socioeconomic or lifestyle factors on IC.

## 2. Methods

### Study cohort: The Canadian Longitudinal Study on Aging (CLSA)

This study utilized data from the CLSA, the largest nationally representative cohort of ageing adults in Canada. The CLSA includes over 50,000 participants aged 45-85 years at baseline, recruited from various regions across Canada, encompassing both urban and rural settings[11, 12]. The cohort is divided into two subgroups: the Tracking Cohort and the Comprehensive Cohort. For tracking cohort data were collected through telephone interviews across the 10 provinces of Canada, whereas the comprehensive cohort participants underwent in-person physical assessments and provided blood and urine samples at 11 data collection sites. For this analysis, we focused on the baseline socioeconomic and lifestyle variables of the comprehensive cohort, which comprises more than 30,000 community-dwelling adults who underwent in-depth physical, clinical, and biological assessments. Of these, genetic data were available for 26,622 participants [13]. After quality control procedures, a final sample of 13,112 individuals with complete data on IC variables and genetic information was retained for analysis.

#### Outcome variable (IC score)

A factor analysis approach was employed to construct the IC score using 14 baseline variables, following a methodology consistent with our previous work in the UK Biobank [7] and this has been previously described [8]. The resulting score demonstrated strong construct and predictive validity, showing expected associations with key health outcomes (e.g., Age, sex, and mortality) and effect directions consistent with prior evidence [8].

### Socioeconomic and lifestyle factors

The socioeconomic variables included educational attainment (assessed by graduation from high school, highest degree achieved, and highest elementary or high school completed), employment history (working status – yes/no at the time of interview), income (assessed by total annual personal income in Canadian dollars – 5 categories), and social engagement (Playing games and musical instruments/singing), and the lifestyle variables were diet, physical activity, smoking, and sleep health. The complete list of variables included in the analysis can be found in Tables 1 and 2. These factors were selected based on our previous literature review, which outlined the list of socioeconomic and lifestyle factors strongly associated with a wide range of health and functional ability domains [10]. To evaluate the cumulative and broader effects of individual lifestyle factors, we computed composite measures of diet and physical activity (specifically, the prospective urban rural epidemiological study (PURE) Healthy Diet Score, the Mediterranean Diet Score, and the Physical Activity Scale for the Elderly (PASE)) and assessed their potential associations with IC [14–17].

**Table 1:** Association of IC with Socioeconomic factors and their interaction with PGS, in CLSA.

| Variable/category | Categories | Main effect: $\beta$ (95% CI) | Interaction effect: $\beta$ (95% CI) |
| --- | --- | --- | --- |
| **Education** |  |  |  |
| **Graduated from high school** | Graduated vs Not | 0.255* (0.180, 0.329) | -0.065 (-0.139, 0.009) |
| **Highest degree achieved** | Trade certificate/diploma vs No Post.sec | -0.146* (-0.211, -0.081) | -0.029 (-0.095, 0.037) |
|  | Non-uni. certificate/diploma vs No Post.sec | -0.017 (-0.076, 0.042) | -0.019 (-0.077, 0.039) |
|  | University certificate vs No Post.sec | 0.062 (-0.020, 0.143) | -0.081 (-0.160, -0.003) |
|  | Bachelor's degree vs No Post.sec | 0.225* (0.169, 0.282) | -0.033 (-0.089, 0.022) |
|  | Uni. degree/cert. above BA vs No Post.sec | 0.340* (0.283, 0.397) | -0.046 (-0.102, 0.010) |
| **Highest elementary or high school completed** | Grade 9 - 10 vs. Grade 8 or lower | 0.138 (-0.005, 0.281) | -0.074 (-0.214, 0.066) |
|  | Grade 11 – 13 vs. Grade 8 or lower | 0.680* (0.557, 0.803) | -0.004 (-0.125, 0.117) |
| **Employment and income** |  |  |  |
| **Currently working** | Working vs Not working | 0.188* (0.121, 0.255) | 0.018 (-0.045, 0.081) |
| **Total personal income** | \$20,000 - \$49,999 vs. Less than \$20,000 | 0.157* (0.111, 0.202) | 0.022 (-0.023, 0.066) |
| | \$50,000 - \$99,999 vs. Less than \$20,000 | 0.253* (0.207, 0.299) | 0.014 (-0.030, 0.058) |
| | \$100,000 - \$149,999 vs. Less than \$20,000 | 0.358* (0.298, 0.418) | 0.014 (-0.044, 0.071) |
| | \$150,000 + vs. Less than \$20,000 | 0.392* (0.322, 0.461) | 0.042 (-0.024, 0.108) |
| **Engagement - social/leisure activity** |  |  |  |
| **Play games, cards, etc** | Several times a year vs. once a year or less | 0.095* (0.045, 0.145) | -0.018 (-0.068, 0.032) |
|  | Several times a month vs. once a year or less | 0.068* (0.020, 0.115) | -0.023 (-0.069, 0.024) |
|  | Several times a week vs. once a year or less | 0.097* (0.054, 0.141) | -0.007 (-0.051, 0.036) |
|  | Every day vs. once a year or less | 0.122* (0.083, 0.161) | -0.023 (-0.061, 0.015) |
| **Play a musical instrument or sing** | Several times a year vs. once a year or less | 0.181* (0.114, 0.249) | -0.010 (-0.074, 0.055) |
|  | Several times a month vs. once a year or less | 0.188* (0.131, 0.244) | -0.021 (-0.076, 0.034) |
|  | Several times a week vs. once a year or less | 0.246* (0.194, 0.298) | -0.020 (-0.072, 0.032) |
|  | Every day vs. once a year or less | 0.283* (0.211, 0.355) | 0.006 (-0.065, 0.076) |

\*Statistically significant associations ( $P_{\text{FDR-adjusted}} < 0.05$ )

**Table 2:**
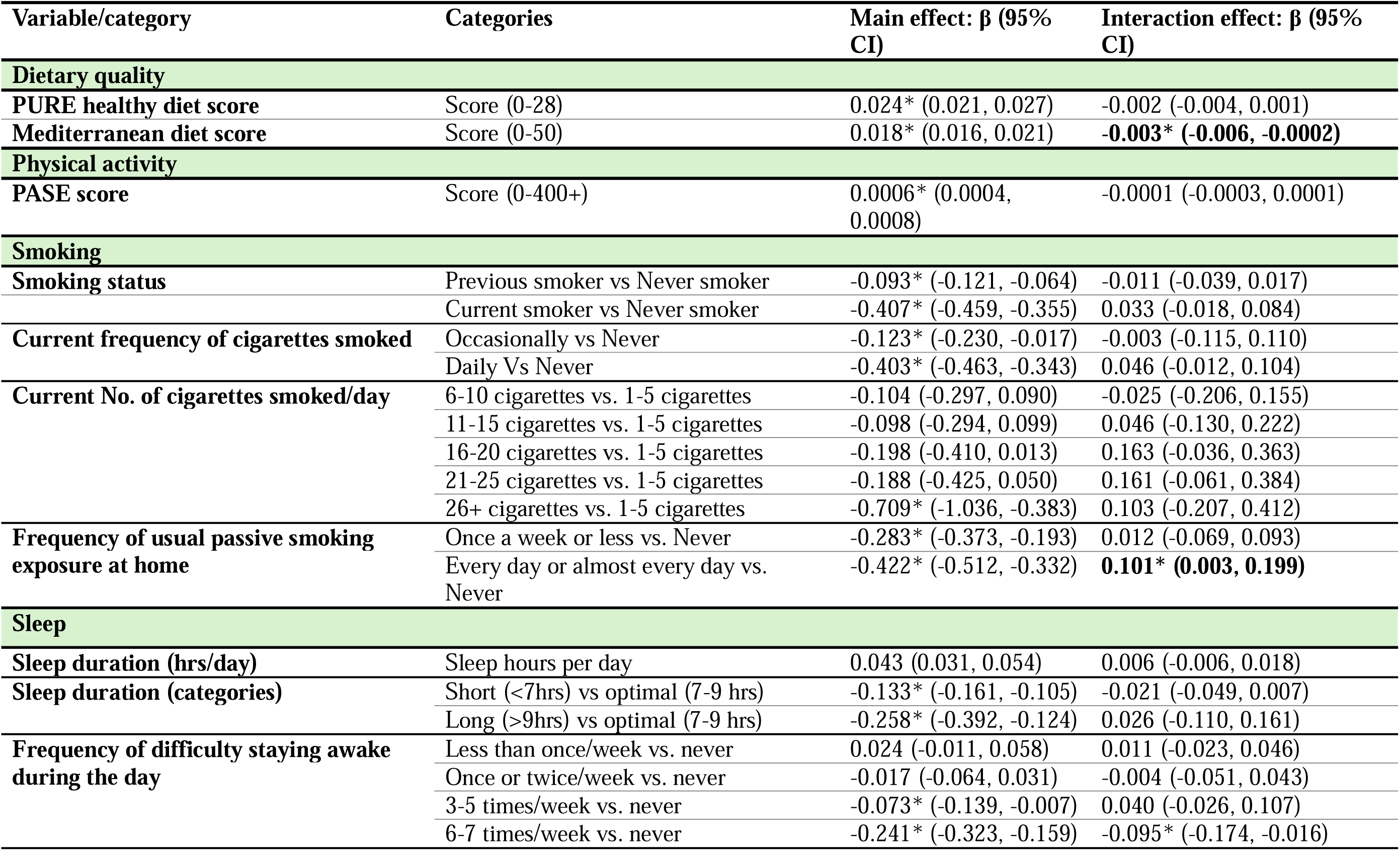
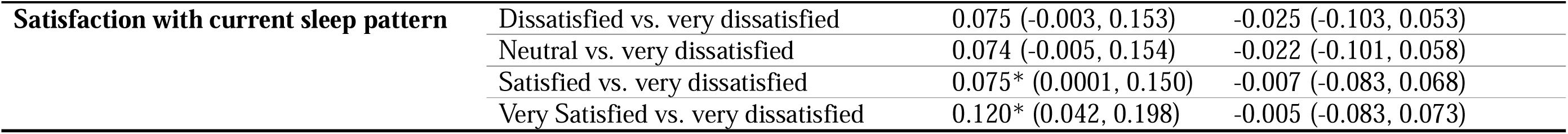
Association of IC with lifestyle factors and their interaction with PGS in the CLSA cohort.

| Variable/category | Categories | Main effect: $\beta$ (95% CI) | Interaction effect: $\beta$ (95% CI) |
| --- | --- | --- | --- |
| **Dietary quality** |  |  |  |
| PURE healthy diet score | Score (0-28) | 0.024* (0.021, 0.027) | -0.002 (-0.004, 0.001) |
| Mediterranean diet score | Score (0-50) | 0.018* (0.016, 0.021) | **-0.003\* (-0.006, -0.0002)** |
| **Physical activity** |  |  |  |
| PASE score | Score (0-400+) | 0.0006* (0.0004, 0.0008) | -0.0001 (-0.0003, 0.0001) |
| **Smoking** |  |  |  |
| Smoking status | Previous smoker vs Never smoker | -0.093* (-0.121, -0.064) | -0.011 (-0.039, 0.017) |
|  | Current smoker vs Never smoker | -0.407* (-0.459, -0.355) | 0.033 (-0.018, 0.084) |
| Current frequency of cigarettes smoked | Occasionally vs Never | -0.123* (-0.230, -0.017) | -0.003 (-0.115, 0.110) |
|  | Daily Vs Never | -0.403* (-0.463, -0.343) | 0.046 (-0.012, 0.104) |
| Current No. of cigarettes smoked/day | 6-10 cigarettes vs. 1-5 cigarettes | -0.104 (-0.297, 0.090) | -0.025 (-0.206, 0.155) |
|  | 11-15 cigarettes vs. 1-5 cigarettes | -0.098 (-0.294, 0.099) | 0.046 (-0.130, 0.222) |
|  | 16-20 cigarettes vs. 1-5 cigarettes | -0.198 (-0.410, 0.013) | 0.163 (-0.036, 0.363) |
|  | 21-25 cigarettes vs. 1-5 cigarettes | -0.188 (-0.425, 0.050) | 0.161 (-0.061, 0.384) |
|  | 26+ cigarettes vs. 1-5 cigarettes | -0.709* (-1.036, -0.383) | 0.103 (-0.207, 0.412) |
| Frequency of usual passive smoking exposure at home | Once a week or less vs. Never | -0.283* (-0.373, -0.193) | 0.012 (-0.069, 0.093) |
|  | Every day or almost every day vs. Never | -0.422* (-0.512, -0.332) | **0.101\* (0.003, 0.199)** |
| **Sleep** |  |  |  |
| Sleep duration (hrs/day) | Sleep hours per day | 0.043 (0.031, 0.054) | 0.006 (-0.006, 0.018) |
| Sleep duration (categories) | Short (<7hrs) vs optimal (7-9 hrs) | -0.133* (-0.161, -0.105) | -0.021 (-0.049, 0.007) |
|  | Long (>9hrs) vs optimal (7-9 hrs) | -0.258* (-0.392, -0.124) | 0.026 (-0.110, 0.161) |
| Frequency of difficulty staying awake during the day | Less than once/week vs. never | 0.024 (-0.011, 0.058) | 0.011 (-0.023, 0.046) |
|  | Once or twice/week vs. never | -0.017 (-0.064, 0.031) | -0.004 (-0.051, 0.043) |
|  | 3-5 times/week vs. never | -0.073* (-0.139, -0.007) | 0.040 (-0.026, 0.107) |
|  | 6-7 times/week vs. never | -0.241* (-0.323, -0.159) | -0.095* (-0.174, -0.016) |
| **Satisfaction with current sleep pattern** | Dissatisfied vs. very dissatisfied | 0.075 (-0.003, 0.153) | -0.025 (-0.103, 0.053) |
|  | Neutral vs. very dissatisfied | 0.074 (-0.005, 0.154) | -0.022 (-0.101, 0.058) |
|  | Satisfied vs. very dissatisfied | 0.075* (0.0001, 0.150) | -0.007 (-0.083, 0.068) |
|  | Very Satisfied vs. very dissatisfied | 0.120* (0.042, 0.198) | -0.005 (-0.083, 0.073) |

#### Diet scores

Nutritional assessment in the CLSA was conducted using the Short Diet Questionnaire (SDQ), a validated tool designed to provide an overview of dietary habits (quantity and frequency of consumption) of the study participants [18]. Using this data, we constructed two widely used and validated composite dietary indices: the PURE Healthy Diet Score [15] and the Mediterranean Diet Score [19].

The *PURE Healthy Diet Score* was calculated based on the frequency of consumption (converted to the number of times per day) of seven healthy food groups: fruits, vegetables, legumes, nuts, fish, dairy, and meats (including red meat, pork, and poultry). Each food group was scored on a quintile-based scale ranging from 0 to 4, with higher scores indicating healthier intake levels. The final score, obtained by summing the scores across all seven groups, ranged from 0 to 28, representing the spectrum from poorest to best diet quality[15, 20]. Detailed components of the PURE Healthy Diet Score are provided in the supplementary methods, section A. Similarly, *the Mediterranean diet score* was calculated using the frequency of consumption (converted to how many times per day) for ten food/drink groups based on CLSA SDQ data [19, 21]. Beneficial components (whole grains, fruits, vegetables, legumes, and nuts, potatoes, and fish) were positively scored, while detrimental components (meat and meat products, poultry, full-fat dairy, and alcohol) were inversely scored. Each group contributed 0 to 5 points, yielding a total score from 0 to 50, with higher scores indicating greater adherence to the Mediterranean diet [19, 21]. Full scoring details are provided in Supplementary Methods, section B.

The ***Physical Activity Scale for the Elderly (PASE)*** is a validated tool for evaluating the frequency and intensity of physical activities in older adults [22]. The total PASE score (0-400+) was derived as the sum of weighted scores from 12 self-reported items covering leisure, household, and occupational activities over the past seven days, based on the original PASE scoring manual [22–24]. Full details of scoring procedures and variable derivations are provided in the Supplementary Methods, section C.

#### Sleep

Sleep duration was assessed based on participants’ self-reported average hours of sleep in a 24-hour period. Later, we categorized them into three groups: short sleep (<7 hours), optimal/recommended sleep (7-9 hours), and long sleep (>9 hours), in line with established sleep health guidelines [25]. In addition, we have also tested trouble falling asleep, difficulty staying awake during the day, as well as satisfaction with sleep patterns.

### Polygenic Score for IC

PGS-IC was developed in our previous study [8] for CLSA participants (target cohort) using the Polygenic Risk Score-Continuous Shrinkage (PRS-CS) method [26], which has been shown to yield higher predictive performance compared to other approaches[27–29]. This method applies a Bayesian regression framework with continuous shrinkage priors to estimate the posterior effect sizes of single-nucleotide polymorphisms (SNPs). The scores were computed using GWAS summary statistics from the UK Biobank (discovery dataset) [8] and linkage disequilibrium (LD) reference data from the European panel of the 1000 Genomes Project [30] to match the predominantly European ancestry of the CLSA sample (97% for our sample) and minimize LD mismatch. A total of 652,994 SNPs were included in the PGS calculation.

### Statistical Analysis

This study employed three independent linear regression models to examine the association of IC with: i) socioeconomic or lifestyle factors, adjusting for age and sex (Model 1); ii) PGS-IC adjusting for age, sex, and the first 5 genetic principal components (PCs) (Model 2) followed by stratification by age groups 45-64 and 65+; and iii) the interaction of PGS and socioeconomic/lifestyle factors, adjusting for the effect of each socioeconomic/lifestyle factor, the PGS-IC, age, sex, and the first 5 PCs (Model 3). The interaction effect analyses were undertaken for (1) the entire sample and (2) with the sample stratified by two age groups: 45-64 and 65+, because there was a differing pattern in the association of PGS-IC with the IC score. For models with significant interaction effects (either in the whole sample or age-stratified analysis), separate linear regression analyses were done to compare the effect of the corresponding socioeconomic or lifestyle factor in individuals with low (decile 1), middle (deciles 2-9), or high (decile 10) PGS-IC.

All analyses were conducted using *lm* function from the “*stats*” package in R (version 4.3.1). Multiple testing corrections were applied using the Benjamini-Hochberg False Discovery Rate (FDR-BH) procedure. Associations were considered statistically significant at an FDR-adjusted p-value < 0.05.

**Model 1**: *IC* = β□ + β□* *Factor_i_* + β□\**Age* + β□\**Sex*+ε*i*

**Model 2**: *IC* = β□ + β□\**PGS* + β□\**Age* + β□\**Sex* + β□\**PC1* + *γ^f^ PC_1-5_* +ε*i*

**Model 3**: *IC* = β□ + β□\**PGS* + β*_i_*\**Factor_i_*+ β□\**(PGS* × *Factor_i_)* + β□\**Age* + β□\**Sex* + *γ^f^PC_1-5_* +ε*i*

Where β *=* betta coefficient, y^f^*PC_1-5_*is a vector of the first five genetic principal components, and ε*i* are error terms.

## 3. Results

The mean (SD) age of the sample was 61 (9.6) years, and 50.8% of participants were females. The cohort had predominantly European (97.4%) genetic ancestry, with smaller representations of Asian (1.4%), African (0.6%), Hispanic (0.4%), and other genetic backgrounds (0.2%). The description of sociodemographic characteristics is detailed in Supplementary Table 1.

### The association of IC with socioeconomic and lifestyle factors

#### Socioeconomic Factors

Among the socioeconomic factors, current employment status, educational attainment, personal income, and participation in leisure activities, such as playing games and music, were significantly associated with IC. For example, participants who reported working during the baseline data collection had higher IC scores compared to those who were not (β = 0.188, 95% CI: 0.121 to 0.255). Educational attainment assessed in the highest school completed, graduation from high school, as well as the highest degree achieved, all showed consistent positive associations with IC (Table 1), with higher levels of education being associated with higher IC scores. For example, individuals who had graduated from high school have higher IC scores than those who had not (β =0.255, 95% CI: 0.180, 0.329). Total personal income also showed a consistent positive association with IC, with all higher income categories demonstrating significantly higher IC scores compared with those in the lowest income group. Similarly, engagement in leisure activities, such as playing games and musical instruments, was positively associated with IC, with greater frequency of participation relating to higher IC scores (Table 1).

#### Diet types and composite diet scores

Dietary intake was significantly associated with IC, with detailed results for each of the 36 food types provided in Supplementary Table 2. Moreover, the two composite healthy diet scores assessed showed strong associations with IC. A higher PURE healthy diet score (β = 0.024, 95% CI: 0.021, 0.027) and greater adherence to the Mediterranean diet (β = 0.018, 95% CI: 0.016, 0.022) were both associated with higher IC levels. Decile-based stratified analyses support these findings with a clear dose-response relationship where IC scores increased consistently across increasing deciles of both diet scores (Figure 1). Detailed results for the association of lifestyle factors with IC are provided in Table 2.

**Figure 1:**
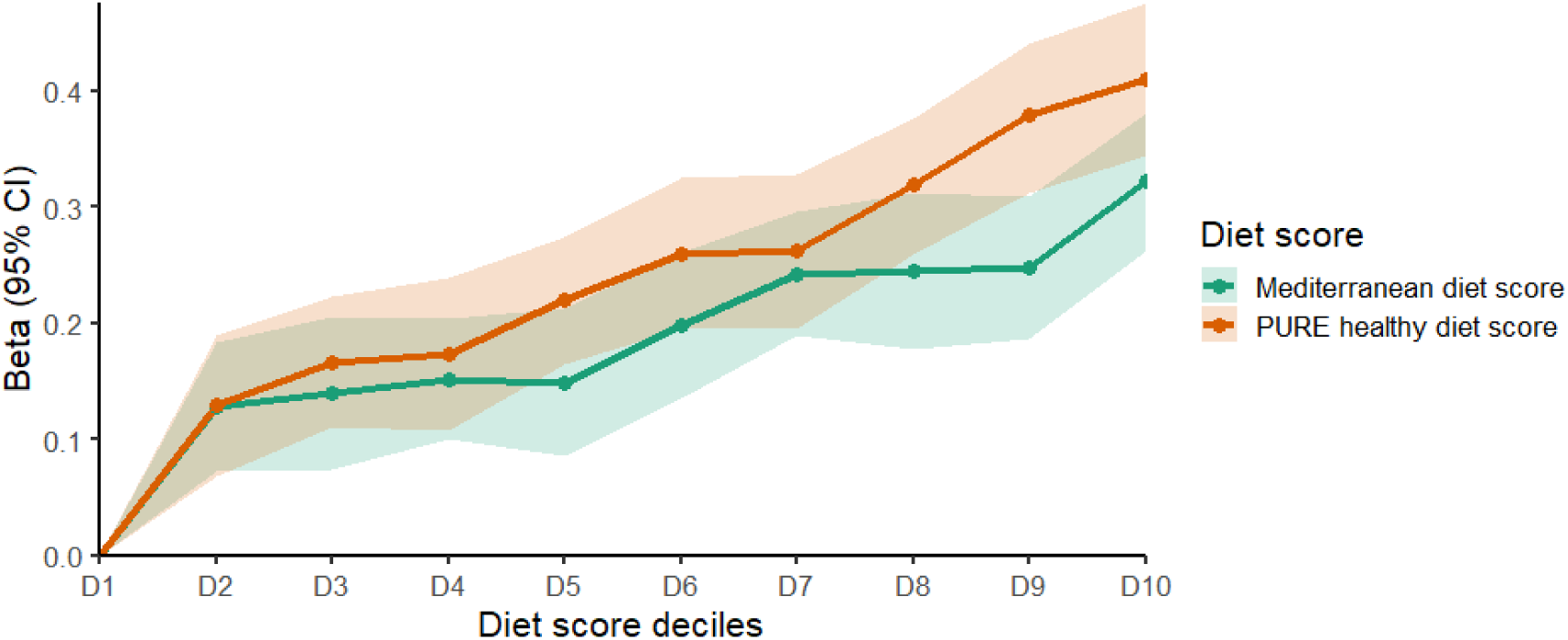
Association between IC and healthy diet scores (across deciles) in the CLSA cohort. Beta coefficients (with 95% confidence intervals) from linear regression models are plotted across deciles (D2-D10) of each diet score, using D1 (individuals with poor healthy diet habits) as the reference category.

#### Cigarette smoking

In this analysis, all the cigarette-smoking-related variables (smoking status, number of cigarettes smoked, and frequency of smoking) showed consistent negative associations with IC.

Current smokers (β = -0.407, 95% CI: -0.459, -0.355) and former smokers (β = -0.093, 95% CI: -0.121, -0.064) had significantly lower IC scores compared to never smokers (Table 2). A similar pattern was observed across smoking frequency: individuals who smoked occasionally had lower IC than never smokers (β = -0.123, 95% CI: -0.230, -0.017), and daily smokers had even lower IC scores than never smokers (β = -0.403, 95% CI: -0.463, -0.343). Regarding the effect of the number of cigarettes smoked per day, a significant inverse association with IC was observed. Specifically, those who smoked 26+ cigarettes per day had a statistically significantly lower IC score compared to those who smoked 1-5 cigarettes a day (β = -0.770, 95% CI: -1.106, -0.435). Passive smoking at home was inversely associated with IC: participants exposed once a week or less had lower IC than those never exposed (β = −0.283; 95% CI: −0.373, −0.193), and those exposed daily or almost daily had even lower IC (β = −0.422; 95% CI: −0.512, −0.332). Additional details in Table 2 and Supplementary Figure 1.

#### Sleep duration and quality

Several sleep-related variables had significant associations with IC. In general, longer average daily sleep duration (assessed as a continuous variable in hours per day) was positively associated with IC (β = 0.043, 95% CI: 0.031, 0.054). Compared to individuals who reported sleeping within the recommended 7 -9 hours per night, those sleeping less than 7 hours had significantly lower IC scores (β = -0.133, 95% CI: -0.161, - 0.105), while those sleeping more than 9 hours had even lower IC scores (β = -0.258, 95% CI: -0.392, -0.124) (Supplementary Figure 2). Individuals who had a higher frequency of daytime drowsiness had, on average, lower IC scores compared to those who never experienced such difficulty (details in Table 2). Satisfaction with current sleep patterns was also positively associated with IC: individuals who reported being very satisfied with their sleep had, on average, 0.12 higher IC scores than those who were very dissatisfied (β = 0.012, 95% CI: 0.042, 0.198).

#### Physical activity

Physical activity, measured using the PASE score, was positively associated with IC. Higher total PASE scores were significantly associated with higher IC scores (β = 0.0006, 95% CI: 0.0004, 0.0008). The decile-based analysis of PASE scores demonstrated a clear dose-response relationship, with individuals in the 2^nd^ to 10^th^ deciles having significantly higher IC scores compared to those in the first decile (with the lowest physical activity score) (Figure 2). The association was strongest in the middle to upper deciles, suggesting that even moderate levels of physical activity may contribute significantly to higher IC.

**Figure 2:**
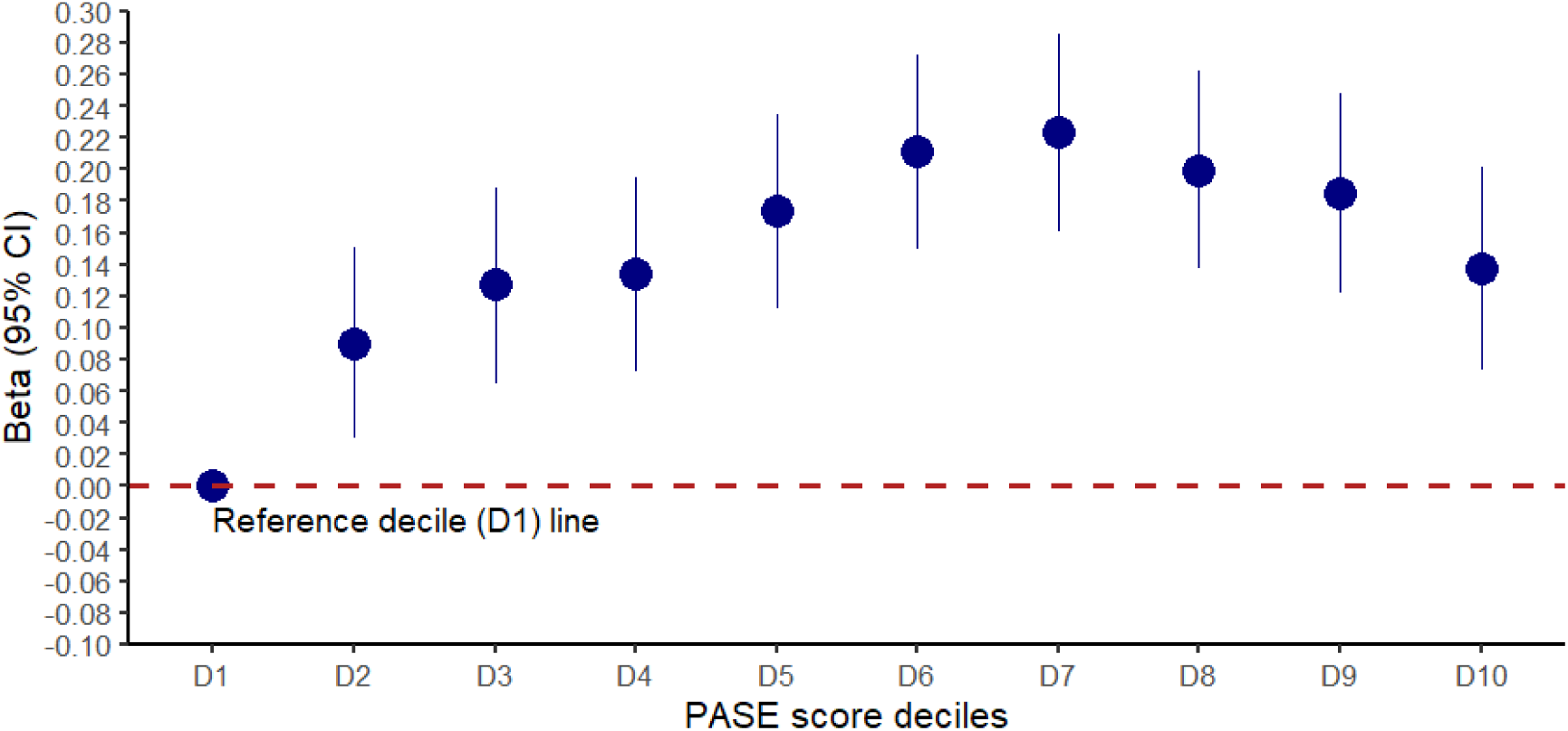
Association between physical activity (measured by total PASE scale, deciles) and IC. This figure displays the beta coefficients and 95% confidence intervals for the association between PASE score deciles and IC. The x-axis represents deciles of the Physical Activity Scale for the Elderly (PASE) score, and the y-axis indicates the estimated beta coefficients for the association of IC with each decile compared to the reference decile. The reference group is the first decile (D1), indicated by the horizontal dashed line at zero.

#### Polygenic scores

The PGS-IC was associated with higher IC, with progressively greater β coefficients observed across higher deciles (D_2_-D_9_) relative to the reference decile, D_1_ (Figure 3). In stratified analyses by age group, we found that the associations were most pronounced in the 45-64 years old age group but weaker and not statistically significant in the ≥ 65 years old age group, with confidence intervals for all deciles reaching or crossing zero (Figure 3).

**Figure 3.**
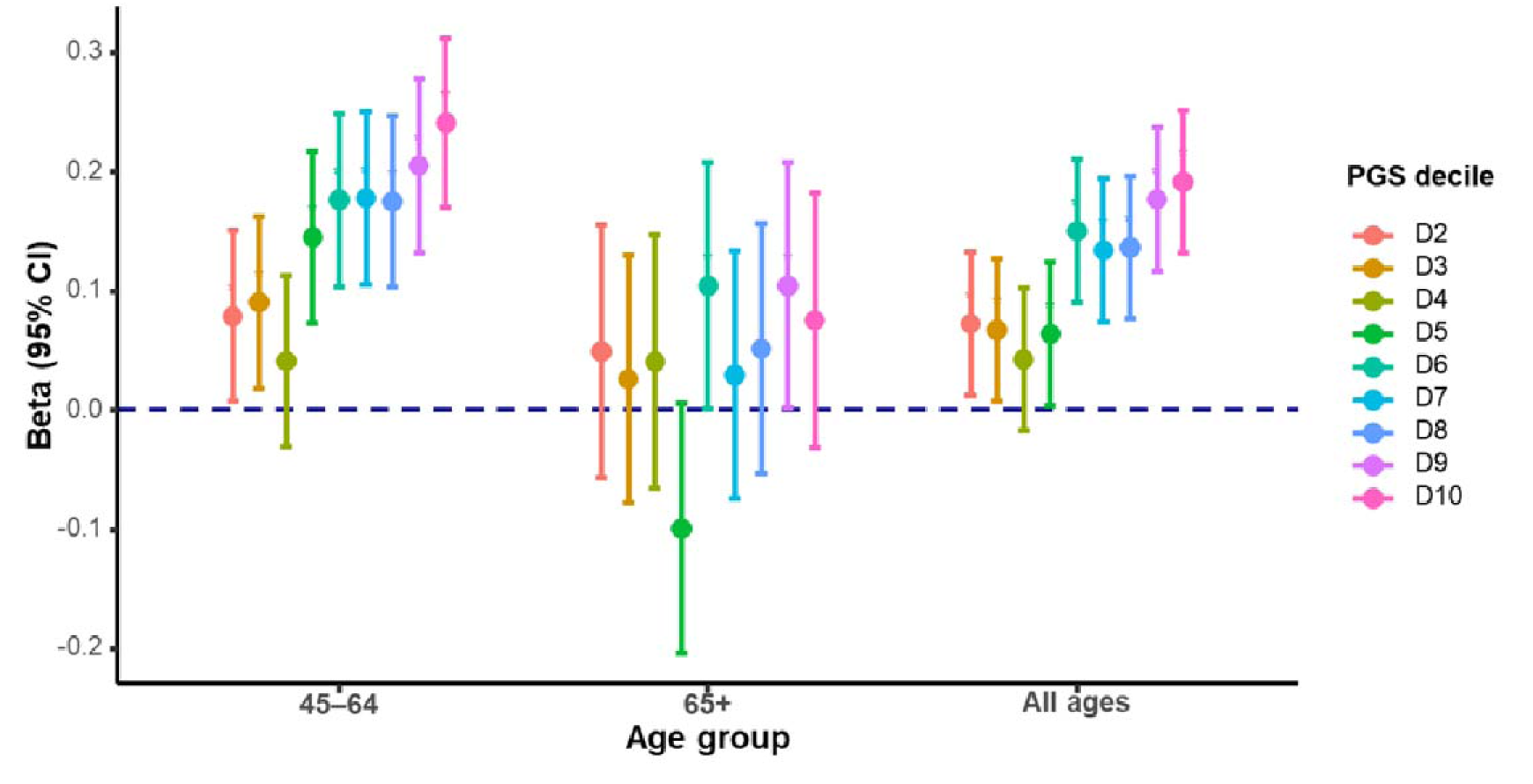
Association of PGS for IC with IC score, stratified by age group. Shown in the figure are linear regression coefficients (β) with 95% confidence intervals for the association of PGS (D_2_ - D_10_) compared with reference, dashed line at β = 0 with IC, separately for participants aged 45 -64 years, ≥65 years, and for the overall sample.

#### Socioeconomic or lifestyle factors and PGS interaction

Our analysis revealed statistically significant interaction effects on IC of the socioeconomic or lifestyle factors and PGS-IC. Specifically, the effects of adherence to the Mediterranean diet (β = -0.003, 95% CI: -0.006 to -0.0002), education - high school completion (β = -0.109, 95% CI: -0.211 to -0.007), and long sleep duration in adults aged 45-64 years (β = 0.198, 95% CI: 0.023 to 0.373), as well as short sleep duration in individuals aged 65 years and older (β = -0.095, 95% CI: -0.153 to - 0.036), were modulated by polygenic loading for IC. These results are provided in Supplementary Table 3. When the effects of these factors on IC were compared across individuals in the low, middle, and high deciles of PGS-IC, the confidence intervals for the effect estimates were overlapping across the PGS-IC categories. Details of the effect estimates of these factors on IC across the PGS-IC categories are presented in the Supplementary Table 4 and Supplementary Figures 3-5.

## Discussion

The novel findings from this study were that significant interaction effects were observed with diet (i.e., adherence to the Mediterranean diet) when the whole sample was analysed, as well as with educational attainment and long sleep duration in younger adults (45-64 years old) and short sleep duration in older adults (65+). Novel associations were also demonstrated in relation to the association of the PASE, PURE, and Mediterranean diet scores with IC. While our earlier study demonstrated an overall association between PGS-IC and IC [8], the current analysis shows that the effect is evident in younger adults, whereas in older adults, it does not reach statistical significance. Our study also reaffirmed associations of IC with key socioeconomic factors, such as education, income, employment, and social engagement, and lifestyle factors such as smoking and sleep health.

Age-dependent pattern of PGS-IC associations with IC, whereby associations are evident in younger adults but not in older ones, may suggest that genetic predisposition plays a stronger role earlier in life, whereas in older age, prolonged exposure to lifestyle factors and related conditions exerts greater influence. This aligns with prior evidence on related traits such as functional ability and cognitive traits, where genetic effects have been shown to decline with age [31, 32]. Such a reduction may mainly reflect age-related alterations in gene expression and genetic effects across the lifespan [32, 33].

The interaction between adherence to a Mediterranean-type diet and PGS-IC indicated greater dietary benefit for individuals with lower genetic loading for IC. This finding points to the potential value of diet as a modifiable factor to help preserve IC, particularly in those with low genetic loading for IC. Comparable evidence has recently been reported for hand grip strength, where leisure-time physical activity exerted a stronger positive effect among individuals with lower PGS for grip strength [34]. In line with this, adherence to the Mediterranean diet has been shown to provide greater benefit for those at high genetic risk of macular degeneration [35]. Similar differential effect of diet with greater benefit for those with high genetic risk had also been reported in the context of other traits. For example, fruit and vegetable intake was found to confer greater protection against obesity among individuals genetically predisposed to higher BMI and weight gain [36].

Education, among younger adults (45-64 years), had a negative interaction effect on IC. i.e., greater effect (benefit) of education for those who had low genetic loading. This is in line with recent evidence, which reported that higher educational attainment attenuates the risk of dementia among those with high genetic predisposition for dementia [37]. Comparable to our finding on sleep duration, a higher benefit of sleep quality (sleep consolidation) on cognitive measures was reported among individuals at higher genetic risk for Alzheimer’s disease [38].

Despite these significant interaction effects detected for diet, sleep, and education, comparisons through separate regression analyses across low, middle, and high PGS groups revealed only suggestive patterns, as confidence intervals overlapped and statistical significance was not reached. This may be due to reduced sample size in the subgroup analysis, which leads to reduced statistical power to detect small effect sizes. For other socioeconomic and lifestyle factors, no evidence of interaction effects on IC of the socioeconomic or lifestyle factors with PGS was observed. These non-significant results may be partly attributable to limited statistical power, as robust detection of PGS × environment interactions typically requires very large sample sizes, a challenge well-documented in recent methodological works [39–42] and observed in studies of cognition [43] and cardiometabolic traits, even across large cohorts [44].

Our association tests with socioeconomic and lifestyle factors reaffirmed the beneficial effects of a healthier lifestyle and higher SES for IC. Among these, physical activity showed a strong positive association with IC, consistent with previous studies [45, 46] and this is the first study to examine this relationship using the PASE scale, a validated tool for older adults. Our findings align with those from large cohort studies such as MAPT and SAGE, which link physical activity to higher baseline IC and slower decline [47, 48], and with intervention trials showing that exercise improves mobility and mental well-being, which are key IC domains [49]. The robust association may reflect the broad physiological benefits of physical activity in promoting healthy longevity by acting on biological ageing processes [50].

Dietary quality was also strongly associated with IC, with a strong dose-dependent pattern showing incremental benefits with increasing PURE and Mediterranean diet scores. These findings add to a growing body of literature highlighting the role of a quality diet in preserving functional ability and healthy longevity [14, 51, 52]. Unlike earlier studies that focused on specific dietary components or pattern types [53, 54], our use of composite indices such as PURE and Mediterranean dietary scores provides a more standardized assessment of dietary quality and its broader impact on IC. The beneficial effects on IC likely act through multiple pathways, including improved metabolic and cardiovascular health [17], reduced systemic inflammation [55], and preservation of cognitive function [56, 57] and biological ageing [16], consistent with multi-omics evidence for the benefits of calorie-restriction in promoting healthy biological ageing [58]. Several clinical trials have also demonstrated that Mediterranean-style diets improve resilience and cognitive functioning [59, 60] and reduce frailty in older populations [51, 61, 62].

Sleep quality and duration were associated with IC: optimal duration (7–9 hours), greater sleep satisfaction, and less daytime sleepiness related to higher IC, consistent with prior works [63–66]. We also reaffirmed inverse associations at both extremes of sleep (>10 h, <6 h) [67].

Our findings about smoking cigarettes showed its detrimental effects on IC; former smokers had lower IC than never smokers, with current smokers having an even lower IC, indicating the benefits of both smoking cessation and avoidance of smoke for preserving IC. These findings align with evidence on the detrimental impact of tobacco on physical and cognitive health [46, 68–71].

Unsurprisingly, socioeconomic status also contributed to variability in IC: higher education and income were associated with higher IC, potentially through their contribution to building cognitive reserve, healthier behaviours, and better access to resources [1, 46, 72–76]. Social engagement, especially cognitively and emotionally stimulating activities such as playing games or music, was likewise related to higher IC, consistent with prior studies demonstrating their impact in protecting against functional decline by enhancing cognitive and emotional resilience [77–79]. Our findings were also in line with the WHO report that health outcomes consistently follow a social gradient, with lower socioeconomic position associated with poorer health across all levels of income [80].

The main strength of this study lies in the novel findings of gene-environment interplay, by computing comprehensive composite measures of diet and a physical activity scale, specifically validated for the older population with IC. However, there are a few limitations to consider while interpreting the findings. First, the CLSA is predominantly of European genetic ancestry, limiting the generalizability of results to other populations. Second, the gene-environment analyses generally demand a large sample size, and we anticipate that we could not detect the interaction effects on IC of some of the lifestyle and socioeconomic factors with PGS-IC due to sample size constraints. Despite these limitations, our work adds to the literature by documenting associations of composite diet and physical-activity measures with IC and by showing that socioeconomic and lifestyle factors exert interaction effects with PGS-IC.

Taken together, our findings underscore that genetic predisposition to IC, particularly in midlife (45–64 years), contributes to individual differences in IC and can buffer or attenuate the effects of socioeconomic and lifestyle factors. Specifically, novel significant interaction effects on IC were detected with education, diet, and sleep health. We also identified strong associations of IC with comprehensive composite measures of diet and physical activity and reaffirmed established links with key socioeconomic factors (education, income, employment, and social engagement) and lifestyle behaviours, notably sleep health and smoking. These results suggest that public health strategies promoting these behaviours are likely to preserve IC and extend healthy longevity. Future work, leveraging larger datasets, should prioritize harmonized measures of IC, longitudinal designs to capture dynamic change in IC, and exploration of biological mediators (in a multi-omics context) that may help explain pathways linking lifestyle, multi-omics factors, and IC.

## Data Availability

All data produced in the present study are available upon reasonable request to the authors

## Acknowledgments

This research was made possible using the data/biospecimens collected by the CLSA. Funding for the CLSA is provided by the Government of Canada through the Canadian Institutes of Health Research (CIHR) under grant reference: LSA 94473 and the Canada Foundation for Innovation, as well as the following provinces, Newfoundland, Nova Scotia, Quebec, Ontario, Manitoba, Alberta, and British Columbia. This research has been conducted using the CLSA dataset [Comprehensive baseline v7.0 and GEN 3 data sets] under Application Number [2304009]. The CLSA is led by Drs. Parminder Raina, Christina Wolfson and Susan Kirkland.

The development, testing, and validation of the Short Diet Questionnaire (SDQ) were carried out among NuAge study participants as part of the Canadian Longitudinal Study on Aging (CLSA) Phase II validation studies, CIHR 2006-2008. The NuAge study was supported by the Canadian Institutes for Health Research (CIHR), Grant number MOP-62842, and the Quebec Network for Research on Aging, a network funded by the Fonds de Recherche du Québec-Santé.

## Declarations

Professor Renuka Visvanathan and Professor John R. Beard are members of the World Health Organisation Clinical Consortium of Healthy Ageing.

The authors declare that the authors alone are responsible for the views expressed in this article and do not necessarily represent the views, decisions, or policies of the institutions with which they are affiliated, nor the views of the CLSA and UKB.

## Data Availability

Data are available from the Canadian Longitudinal Study on Aging (www.clsa-elcv.ca) and the UK Biobank for researchers who meet the criteria for access to de-identified data.

## References

1. Beard, J.R., et al., The World report on ageing and health: a policy framework for healthy ageing. The Lancet, 2016. 387(10033): p. 2145–2154.

2. Cesari, M., et al., Frailty: An Emerging Public Health Priority. Journal of the American Medical Directors Association, 2016. 17(3): p. 188–192.

3. Chatterji, S., et al., Health, functioning, and disability in older adults—present status and future implications. The Lancet, 2015. 385(9967): p. 563–575.

4. Rudnicka, E., et al., The World Health Organization (WHO) approach to healthy ageing. Maturitas, 2020. 139: p. 6–11.

5. World Health, O., World report on ageing and health. 2015, Geneva: World Health Organization.

6. Beard, J.R., et al., The structure and predictive value of intrinsic capacity in a longitudinal study of ageing. BMJ Open, 2019. 9(11): p. e026119.

7. Beyene, M.B., et al., Development and validation of an intrinsic capacity score in the UK Biobank study. Maturitas, 2024. 185: p. 107976.

8. Beyene, M.B., et al., A genome-wide association study identified 10 novel genomic loci associated with intrinsic capacity. medRxiv, 2025: p. 2025.02.05.25321753.

9. Fuentealba, M., et al., A blood-based epigenetic clock for intrinsic capacity predicts mortality and is associated with clinical, immunological and lifestyle factors. Nature Aging, 2025.

10. Beyene, M.B., R. Visvanathan, and A.T. Amare, Intrinsic Capacity and Its Biological Basis: A Scoping Review. The Journal of Frailty & Aging, 2024. 13(3): p. 193–202.

11. Raina, P.S., et al., The Canadian longitudinal study on aging (CLSA). Can J Aging, 2009. 28(3): p. 221–9.

12. Raina, P., et al., Cohort Profile: The Canadian Longitudinal Study on Aging (CLSA). International Journal of Epidemiology, 2019. 48(6): p. 1752–1753j.

13. Forgetta, V., et al., Cohort profile: genomic data for 26 622 individuals from the Canadian Longitudinal Study on Aging (CLSA). BMJ Open, 2022. 12(3): p. e059021.

14. Kant, A.K., Dietary patterns and health outcomes. Journal of the American Dietetic Association, 2004. 104(4): p. 615–635.

15. Mente, A., et al., Diet, cardiovascular disease, and mortality in 80 countries. Eur Heart J, 2023. 44(28): p. 2560–2579.

16. Milte, C.M. and S.A. McNaughton, Dietary patterns and successful ageing: a systematic review. Eur J Nutr, 2016. 55(2): p. 423–450.

17. Papadaki, A., E. Nolen-Doerr, and C.S. Mantzoros, The Effect of the Mediterranean Diet on Metabolic Health: A Systematic Review and Meta-Analysis of Controlled Trials in Adults. Nutrients, 2020. 12(11): p. 3342.

18. Shatenstein, B. and H. Payette, Evaluation of the Relative Validity of the Short Diet Questionnaire for Assessing Usual Consumption Frequencies of Selected Nutrients and Foods. Nutrients, 2015. 7(8): p. 6362–74.

19. Aoun, C., et al., Comparison of five international indices of adherence to the Mediterranean diet among healthy adults: similarities and differences. Nutrition research and practice, 2019. 13(4): p. 333–343.

20. Bassim, C., et al., Oral health, diet, and frailty at baseline of the Canadian longitudinal study on aging. Journal of the American Geriatrics Society, 2020. 68(5): p. 959–966.

21. Vahid, F., P. Wilk, and T. Bohn, Longitudinal effects of diet quality on healthy aging - Focus on cardiometabolic health: findings from the Canadian longitudinal study on aging (CLSA). Aging Clinical and Experimental Research, 2025. 37(1): p. 157.

22. Washburn, R.A., et al., The physical activity scale for the elderly (PASE): Development and evaluation. Journal of Clinical Epidemiology, 1993. 46(2): p. 153–162.

23. Institutes, N.E.R., PASE: Physical Activity Scale for the Elderly: Administration and Scoring Instruction Manual. 1991, New England Research Institutes Watertown, MA.

24. D’Amore, C., et al., Physical Activity Behaviour in Middle-Aged and Older Canadian Women and Men: An Analysis of the CLSA. medRxiv, 2025: p. 2025.03.17.25323990.

25. Watson, N.F., et al., Recommended Amount of Sleep for a Healthy Adult: A Joint Consensus Statement of the American Academy of Sleep Medicine and Sleep Research Society. Sleep, 2015. 38(6): p. 843–4.

26. Ge, T., et al., Polygenic prediction via Bayesian regression and continuous shrinkage priors. Nature Communications, 2019. 10(1): p. 1776.

27. Amare, A.T., et al., Association of polygenic score for major depression with response to lithium in patients with bipolar disorder. Molecular Psychiatry, 2021. 26(6): p. 2457–2470.

28. Amare, A.T., et al., Association of polygenic score and the involvement of cholinergic and glutamatergic pathways with lithium treatment response in patients with bipolar disorder. Molecular Psychiatry, 2023. 28(12): p. 5251–5261.

29. Sharew, N.T., et al., Pathway-Specific Polygenic Scores for Predicting Clinical Lithium Treatment Response in Patients With Bipolar Disorder. Biol Psychiatry Glob Open Sci, 2025. 5(5): p. 100558.

30. Auton, A., et al., A global reference for human genetic variation. Nature, 2015. 526(7571): p. 68–74.

31. Kim, A., et al., The Heritability of Cognitive Aging: A Systematic Review of Longitudinal Twin Studies. Innovation in Aging, 2021. 5(Supplement_1): p. 1017–1017.

32. Ding, X., et al., The relationship between cognitive decline and a genetic predictor of educational attainment. Soc Sci Med, 2019. 239: p. 112549.

33. Viñuela, A., et al., Age-dependent changes in mean and variance of gene expression across tissues in a twin cohort. Hum Mol Genet, 2018. 27(4): p. 732–741.

34. Herranen, P., et al., Evaluating a Genome-Wide Polygenic Score for Handgrip Strength and Its Interplay with Leisure-Time Physical Activity Across the IGEMS Twin Cohorts. medRxiv, 2025: p. 2025.08.07.25333195.

35. Barreto, P., et al., Interaction between genetics and the adherence to the Mediterranean diet: the risk for age-related macular degeneration. Coimbra Eye Study Report 8. Eye and Vision, 2023. 10(1): p. 38.

36. Wang, T., et al., Improving fruit and vegetable intake attenuates the genetic association with long-term weight gain. The American Journal of Clinical Nutrition, 2019. 110(3): p. 759–768.

37. Ma, H., et al., Early-life educational attainment, APOE ε4 alleles, and incident dementia risk in late life. GeroScience, 2022. 44(3): p. 1479–1488.

38. Huang, L.-Y., et al., Sleep quality, APOE ε4, and Alzheimer’s disease: associations from two prospective cohort studies and mechanisms by plasma proteomic analysis. BMC Medicine, 2025. 23(1): p. 462.

39. Duncan, L.E. and M.C. Keller, A critical review of the first 10 years of candidate gene-by-environment interaction research in psychiatry. Am J Psychiatry, 2011. 168(10): p. 1041–9.

40. Dudbridge, F., Power and predictive accuracy of polygenic risk scores. PLoS Genet, 2013. 9(3): p. e1003348.

41. Luan, J., et al., Sample size determination for studies of gene-environment interaction. International Journal of Epidemiology, 2001. 30(5): p. 1035–1040.

42. Plomin, R., et al., Gene-environment interaction using polygenic scores: Do polygenic scores for psychopathology moderate predictions from environmental risk to behavior problems? Dev Psychopathol, 2022. 34(5): p. 1816–1826.

43. von Stumm, S. and A.F. Nancarrow, New methods, persistent issues, and one solution: Gene-environment interaction studies of childhood cognitive development. Intelligence, 2024. 105: p. 101834.

44. Hartiala, J.A., et al., Gene-Environment Interactions for Cardiovascular Disease. Current Atherosclerosis Reports, 2021. 23(12): p. 75.

45. Zhou, M., L. Kuang, and N. Hu, The Association between Physical Activity and Intrinsic Capacity in Chinese Older Adults and Its Connection to Primary Care: China Health and Retirement Longitudinal Study (CHARLS). Int J Environ Res Public Health, 2023. 20(7).

46. Wei, X., et al., Factors associated with the intrinsic capacity in older adults: A scoping review. J Clin Nurs, 2024. 33(5): p. 1739–1750.

47. Raffin, J., et al., Cross-sectional and longitudinal associations between physical activity and intrinsic capacity in healthy older adults from the MAPT study. Archives of Gerontology and Geriatrics, 2025. 130: p. 105724.

48. Kowal, P., et al., Data Resource Profile: The World Health Organization Study on global AGEing and adult health (SAGE). International Journal of Epidemiology, 2012. 41(6): p. 1639–1649.

49. Wang, Z., K. Qi, and P. Zhang, Effect of physical activity interventions on physical and mental health of the elderly: a systematic review and meta-analysis. Aging Clin Exp Res, 2025. 37(1): p. 169.

50. Izquierdo, M., et al., Global consensus on optimal exercise recommendations for enhancing healthy longevity in older adults (ICFSR). The Journal of nutrition, health and aging, 2025. 29(1): p. 100401.

51. Ghosh, T.S., et al., Mediterranean diet intervention alters the gut microbiome in older people reducing frailty and improving health status: the NU-AGE 1-year dietary intervention across five European countries. Gut, 2020. 69(7): p. 1218–1228.

52. Bellavia, A., et al., Quantifying the benefits of Mediterranean diet in terms of survival. Eur J Epidemiol, 2016. 31(5): p. 527–30.

53. Huang, C.H., et al., Dietary patterns and intrinsic capacity among community-dwelling older adults: a 3-year prospective cohort study. Eur J Nutr, 2021. 60(6): p. 3303–3313.

54. Gómez-Cao, M., et al., Association between a Planetary Health Diet and changes in intrinsic capacity in older adults: the Seniors-ENRICA cohorts. Age and Ageing, 2025. 54(6).

55. Tsigalou, C., et al., Mediterranean Diet as a Tool to Combat Inflammation and Chronic Diseases. An Overview. Biomedicines, 2020. 8(7).

56. Chen, X., et al., Dietary Patterns and Cognitive Health in Older Adults: A Systematic Review. J Alzheimers Dis, 2019. 67(2): p. 583–619.

57. Féart, C., et al., Adherence to a Mediterranean diet, cognitive decline, and risk of dementia. Jama, 2009. 302(6): p. 638–48.

58. Fiorito, G., et al., Multi-omic analysis of biological aging biomarkers in long-term calorie restriction and endurance exercise practitioners: A cross-sectional study. Aging Cell, 2025. 24(4): p. e14442.

59. Marseglia, A., et al., Effect of the NU-AGE Diet on Cognitive Functioning in Older Adults: A Randomized Controlled Trial. Front Physiol, 2018. 9: p. 349.

60. Valls-Pedret, C., et al., Mediterranean Diet and Age-Related Cognitive Decline: A Randomized Clinical Trial. JAMA Internal Medicine, 2015. 175(7): p. 1094–1103.

61. Guasch-Ferré, M., et al., The PREDIMED trial, Mediterranean diet and health outcomes: How strong is the evidence? Nutr Metab Cardiovasc Dis, 2017. 27(7): p. 624–632.

62. Zhang, H., et al., Mediterranean diet associated with lower frailty risk: A large cohort study of 21,643 women admitted to hospitals. The Journal of nutrition, health and aging, 2024. 28(1): p. 100001.

63. Chang, Y.H., et al., Association between sleep health and intrinsic capacity among older adults in Taiwan. Sleep Med, 2023. 109: p. 98–103.

64. Zhang, N., et al., Sleep disturbances and intrinsic capacity trajectories among Chinese older adults: The Rugao Longevity and Ageing Study. Geriatric Nursing, 2024. 60: p. 150–155.

65. Chen, X.-L., et al., Validation of intrinsic capacity and healthy sleep pattern in middle-aged and older adults: a longitudinal Chinese study assessing healthy ageing. The Journal of nutrition, health and aging, 2024. 28(11): p. 100365.

66. Cao, Y., et al., Both Short and Long Sleep Durations Are Associated with Poor Cognition and Memory in Chinese Adults Aged 55+ Years-Results from China Health and Nutrition Survey. Life (Basel), 2022. 12(11).

67. Chen, X.L., et al., Validation of intrinsic capacity and healthy sleep pattern in middle-aged and older adults: a longitudinal Chinese study assessing healthy ageing. J Nutr Health Aging, 2024. 28(11): p. 100365.

68. Anstey, K.J., et al., Smoking as a risk factor for dementia and cognitive decline: a meta-analysis of prospective studies. Am J Epidemiol, 2007. 166(4): p. 367–78.

69. Sabia, S., et al., Smoking history and cognitive function in middle age from the Whitehall II study. Arch Intern Med, 2008. 168(11): p. 1165–73.

70. Gellert, C., B. Schöttker, and H. Brenner, Smoking and all-cause mortality in older people: systematic review and meta-analysis. Arch Intern Med, 2012. 172(11): p. 837–44.

71. Oberg, M., et al., Worldwide burden of disease from exposure to second-hand smoke: a retrospective analysis of data from 192 countries. Lancet, 2011. 377(9760): p. 139–46.

72. Marmot, M. and R. Bell, Fair society, healthy lives. Public Health, 2012. 126 Suppl 1: p. S4–s10.

73. Ruiz, M., et al., Life Course Socioeconomic Position and Cognitive Aging Trajectories: A Cross-National Cohort Study in China and England. Innov Aging, 2023. 7(6): p. igad064.

74. Salinas-Rodríguez, A., et al., Intrinsic capacity trajectories and socioeconomic inequalities in health: the contributions of wealth, education, gender, and ethnicity. International Journal for Equity in Health, 2024. 23(1): p. 48.

75. Clouston, S.A.P., et al., Education and Cognitive Decline: An Integrative Analysis of Global Longitudinal Studies of Cognitive Aging. J Gerontol B Psychol Sci Soc Sci, 2020. 75(7): p. e151–e160.

76. Stern, Y., Cognitive reserve in ageing and Alzheimer’s disease. The Lancet Neurology, 2012. 11(11): p. 1006–1012.

77. Mayne, K., et al., Aging and Neurodegenerative Disease: Is the Adaptive Immune System a Friend or Foe? Frontiers in Aging Neuroscience, 2020. **Volume** 12 - 2020.

78. Lieber, S.B., et al., Social support and physical activity: does general health matter? European Review of Aging and Physical Activity, 2024. 21(1): p. 16.

79. Huang, Z.-T., et al., Social determinants of intrinsic capacity: A systematic review of observational studies. Ageing Research Reviews, 2024. 95: p. 102239.

80. Organization, W.H. Social determinants of health. 2025; Available from: https://www.who.int/health-topics/social-determinants-of-health#tab=tab_1.

