## Supplementary Material for "Associations and Interaction Effects of Socioeconomic, Lifestyle, and Genetic Factors on Intrinsic Capacity"

### **Supplementary Methods**

### A. **Prospective Urban Rural Epidemiological (PURE) study healthy diet score**

In this study, we restricted the prospective urban-rural epidemiological (PURE) healthy diet score based on the original PURE diet score [1] and the customized application of the method in the Canadian Longitudinal Study on Aging (CLSA)[2]. As per this validated tool, we included seven healthy diet categories in the calculation of the score. We compute quantiles (0-4) of the daily frequency of intake, with quintile 0 representing the lowest intake and quintile 4 denoting the highest intake of the corresponding food. The total PURE health diet score then is the sum of the quintiles of daily intake frequency for the seven food groups, which yields a score of 0 (worst diet) to 28 (best diet). For food groups where more than one food is included, we add the daily frequency of intake for all included and then compute the quintiles, which then represent the overall intake frequency of the corresponding food group. The details are in the table below.

| Food group | Included foods | Code in CLSA |
| --- | --- | --- |
| Fruits | Fruits | NUT_FRUT_NB_COM |
| Vegetables | Green salads, Carrots, and  Vegetables | NUT_GREEN_NB_COM  NUT_CRRT_NB_COM  NUT_VGOT_NB_COM |
| Legumes | Legumes | NUT_LEGM_NB_COM |
| Nuts | Nuts | NUT_NUTS_NB_COM |
| Fish | Fish | NUT_FISH_NB_COM |
| Dairy | Cheese (regular and low-fat)  Yoghurt (regular and low-fat)  Milk (whole milk, skim milk, and calcium fortified milk intake) | NUT_LWCS_NB_COM  NUT_CHSE_NB_COM  NUT_LWYG_NB_COM  NUT_YOGR_NB_COM  NUT_WHML_NB_COM  NUT_LFML_NB_COM  NUT_CAML_COM |
| Red Meats, pork, and chicken | Beef, pork, veal, lamb, game, and chicken intake | NUT_MEAT_NB_COM  NUT_MTOT_NB_COM  NUT_CHCK_NB_COM |

### B.  **Mediterranean diet score**

The Mediterranean diet score was calculated using dietary intake data from the Short Diet Questionnaire (SDQ) administered in the Canadian Longitudinal Study on Aging (CLSA), based on previously established methods [3, 4]. Reported frequencies of food consumption were converted into estimated times per day. The score was constructed using ten food and beverage groups categorized as either beneficial or detrimental in alignment with the Mediterranean dietary pattern [4].

Beneficial components included whole grains, fruits, vegetables, legumes and nuts, potatoes, and fish. These were scored positively, with higher consumption associated with higher scores. Detrimental components—meat and meat products, poultry, full-fat dairy products, and alcohol—were scored inversely, so that higher consumption resulted in lower scores. Specifically, for alcohol, lower consumption was coded to have a higher score, then no consumption follows, and then the incrementally higher consumption assumes the least values.

Each component contributed 0 to 5 points (which were quintiles of daily frequency of intake), resulting in a total Mediterranean diet score ranging from 0 to 50, with higher scores indicating greater adherence to the Mediterranean dietary pattern. Details of the variables used in the calculation are provided in the following table.

| R. No | Food group | Food items included | Beneficial (+) or detrimental (-) | Codes in CLSA |
| --- | --- | --- | --- | --- |
| 1 | **Whole grains** | High fibre breakfast cereals, whole wheat breads, bran breads, multigrain breads, and ryebreads | **+** | NUT_FBR_NB_COM  NUT_BRD_NB_COM |
| 2 | **Fruits** | Fruits | **+** | NUT_FRUT_NB_COM |
| 3 | **Vegetables** | Green Salads, carrots, and all other vegetables except potatoes | **+** | NUT_GREEN_NB_COM NUT_CRRT_NB_COM NUT_VGOT_NB_COM |
| 4 | **Legumes and Nuts** | Legumes, Nuts | **+** | NUT_LEGM_NB_COM  NUT_NUTS_NB_COM |
| 5 | **Potatoes** | Potatoes | **+** | NUT_PTTO_NB_COM |
| 6 | **Fish** | Fish | **+** | NUT_FISH_NB_COM |
| 7 | **Full-fat dairy** | Cheese, Yoghurt, and whole milk |  | NUT_CHSE_NB_COM NUT_YOGR_NB_COM  NUT_WHML_NB_COM |
| 8 | **Meats** | Beef, Pork, and other meats (veal, lamb, game) |  | NUT_MEAT_NB_COM NUT_MTOT_NB_COM |
| 9 | **Poultry** | Chicken |  | NUT_CHCK_NB_COM |
| 10 | **Alcohol** | Alcohol |  | ALC_FREQ_COM |

### C.  **Physical activity Scale for the elderly (PASE)**

The Physical Activity Scale for the Elderly (PASE) was used to assess participants’ physical activity over the past seven days of the assessment, incorporating leisure, household, and occupational domains. The score was computed from 12 self-reported items using the original PASE scoring manual [5, 6]), with adaptations for the CLSA data structure [7]. For five activities (walking, light sport, moderate sport, strenuous sport, and strength exercise), scores were calculated using frequency × duration × activity-specific weight. Frequency and duration responses were converted as follows:

- **Frequency to days/week**
  - Never = 0
  - Seldom (1–2 days/week) = 1.5
  - Sometimes (3–4 days/week) = 3.5
  - Often (5–7 days/week) = 6
- **Duration in hours/day**
  - Less than 30 minutes = 0.25
  - 30 minutes to 1 hour = 0.75
  - 1–2 hours = 1.5
  - 2–4 hours = 3
  - 4 or more hours = 4

For work-related activity, reported hours per week (capped at 40 hours) were converted to hours per day and scored only if the work involved physical effort (categories 2-4 of the intensity of work question). The remaining six items-housework, home repairs, yard work, outdoor activity, and caregiving-were binary and scored using fixed weights. The total PASE score was the sum of all component scores, with higher values indicating greater physical activity. The list of variables and scoring details is provided in the table below.

| R. No | Physical activity | Variables we used to calculate the score | PASE weight |
| --- | --- | --- | --- |
| 1 | **Walking** | Average hours per day spent walking (PA2_WALKHR_MCQ) and Frequency of taking a walk outside (PA2_WALK_MCQ). | 20 |
| 2 | **Light sports** | Average hours per day engaged in light sports (PA2_LSPRTHR_MCQ) and Frequency of participation in light sports (PA2_LSPRT_MCQ) | 21 |
| 3 | **Moderate sports** | Average hours per day engaged in moderate sports (PA2_MSPRTHR_MCQ) and Frequency of participation in moderate sports (PA2_MSPRT_MCQ) | 23 |
| 4 | **Strenuous sports** | Average hours per day engaged in strenuous sports (PA2_SSPRTHR_MCQ) and Frequency of participation in strenuous sports (PA2_SSPRT_MCQ) | 23 |
| 5 | **Strength exercise** | Average hours per day exercised to increase muscle strength and endurance (PA2_EXERHR_MCQ) and its frequency (PA2_EXER_MCQ) | 30 |
| 6 | **Work involving PA (voluntary or paid)** | Hours worked for pay or as a volunteer (PA2_WRKHRS_NB_MCQ) and the Amount of physical activity required on the job or volunteering (PA2_WRKPA_MCQ) | 21 |
| 7 | **Light housework** | Engaged in light housework (PA2_LTHSWK_MCQ) | 25 |
| 8 | **Heavy housework** | Engaged in heavy housework or chores (PA2_HVYHSWK_MCQ) | 25 |
| 9 | **Home repairs** | Engaged in home repairs (PA2_HMREPAIR_MCQ) | 30 |
| 10 | **Heavy outdoor** | Engaged in lawn work or yard care (PA2_HVYODA_MCQ) | 36 |
| 11 | **Light outdoor** | Engaged in outdoor gardening, sweeping the balcony or the stairs (PA2_LTODA_MCQ) | 20 |
| 12 | **Care giving** | Engaged in caring for another person (PA2_CRPRSN_MCQ) | 35 |

**Supplementary Table 1**: Sociodemographic characteristics of the study sample, CLSA (N= 13,1112)

| Characteristics | Categories | N (%) |
| --- | --- | --- |
| Age in years | [45-53) | 3283 (25.04) |
|  | [53-60) | 3322 (25.33) |
|  | [60-68) | 3461 (26.40) |
|  | [68-86) | 3046 (23.23) |
| Sex | Male | 6453 (49.21) |
|  | Female | 6659 (50.79) |
| Education | < 2^0^ school graduation | 471 (3.40) |
|  | 2^0^, no post-sec. | 1092 (8.34) |
|  | Some post-2^0^ education | 893 (6.82) |
|  | Post-2^0^ degree/diploma | 10635(81.24) |
| Ethnicity | Caucasian | 12333 (97.43) |
|  | Asian | 174 (1.38) |
|  | African | 79 (0.62) |
|  | Hispanic | 44 (0.35) |
|  | Other ethnicity | 28 (0.22) |
| Country of birth | Canada | 10856 (82.80) |
|  | Others | 2255 (17.20) |
| Personal income/year | < $20,000 | 1552 (12.35) |
|  | [$20,000, $50,000) | 4330 (34.44) |
|  | [$50,000, $100,000) | 4607 (36.65) |
|  | [$100,000, $150,000) | 1294 (10.29) |
|  | >$150,000 | 787 (6.26) |
| Marital status | Single | 975 (7.55) |
|  | Married/Living with a partner | 9401 (72.82) |
|  | Widowed | 903 (6.99) |
|  | Divorced | 1278 (9.90) |
|  | Separated | 354 (2.74) |

**Supplementary Table 2:** Association of individual dietary intake with IC**,** in CLSA

| Diet type or composite | Category / Measurement | Beta (95% CI) | P_Overall | P_Category |
| --- | --- | --- | --- | --- |
| Pure fruit juice intake | 1.rarely/never | -0.007 (-0.016, 0.001) | 0.097046038 | 0.097046038 |
|  | 2. Per Year |  |  |  |
|  | 2. Per Month |  |  |  |
|  | 3. Per week |  |  |  |
|  | 5. Per day |  |  |  |
| Skim Milk intake | 1.rarely/never | 0.002 (-0.006,0.010) | 0.663144603 | 0.663144603 |
|  | 2. Per Year |  |  |  |
|  | 2. Per Month |  |  |  |
|  | 3. Per week |  |  |  |
|  | 5. Per day |  |  |  |
| All other egg intake (except Omega-3 eggs) | 1.rarely/never | -0.016 (-0.028, -0.004) | 0.007411893 | 0.007411893 |
|  | 2. Per Year |  |  |  |
|  | 2. Per Month |  |  |  |
|  | 3. Per week |  |  |  |
|  | 5. Per day |  |  |  |
| Low-fat Cheese intake | 1.rarely/never | 0.023 (0.013, 0.033) | 1.20E-05 | 1.20E-05 |
|  | 2. Per Year |  |  |  |
|  | 2. Per Month |  |  |  |
|  | 3. Per week |  |  |  |
|  | 5. Per day |  |  |  |
| Regular Cheese intake | 1.rarely/never | 0.011 (-0.002, 0.024) | 0.093643472 | 0.093643472 |
|  | 2. Per Year |  |  |  |
|  | 2. Per Month |  |  |  |
|  | 3. Per week |  |  |  |
|  | 5. Per day |  |  |  |
| Butter or Regular Margarine intake | 1.rarely/never | -0.019 (-0.029, -0.0099) | 0.000146118 | 0.000146118 |
|  | 2. Per Year |  |  |  |
|  | 2. Per Month |  |  |  |
|  | 3. Per week |  |  |  |
|  | 5. Per day |  |  |  |
| Calcium fortified food intake | 1.rarely/never | 0.03 (0.011, 0.048) | 0.002073767 | 0.002073767 |
|  | 2. Per Year |  |  |  |
|  | 2. Per Month |  |  |  |
|  | 3. Per week |  |  |  |
|  | 5. Per day |  |  |  |
| Calcium-fortified juice intake | 1.rarely/never | -0.022 (-0.038, -0.007) | 0.005105052 | 0.005105052 |
|  | 2. Per Year |  |  |  |
|  | 2. Per Month |  |  |  |
|  | 3. Per week |  |  |  |
|  | 5. Per day |  |  |  |
| Calcium fortified milk intake | 1.rarely/never | -0.023 (-0.058, 0.011) | 0.176350684 | 0.176350684 |
|  | 2. Per Year |  |  |  |
|  | 2. Per Month |  |  |  |
|  | 3. Per week |  |  |  |
|  | 5. Per day |  |  |  |
| Carrots intake | 1.rarely/never | 0.057 (0.036, 0.079) | 1.77E-07 | 1.77E-07 |
|  | 2. Per Year |  |  |  |
|  | 2. Per Month |  |  |  |
|  | 3. Per week |  |  |  |
|  | 5. Per day |  |  |  |
| Chicken intake | 1.rarely/never | -0.049 (-0.073, -0.025) | 0.000147626 | 0.000147626 |
|  | 2. Per Year |  |  |  |
|  | 2. Per Month |  |  |  |
|  | 3. Per week |  |  |  |
|  | 5. Per day |  |  |  |
| Chocolate bar intake | 1.rarely/never | 0.008 (-0.002, 0.019) | 0.129140643 | 0.129140643 |
|  | 2. Per Year |  |  |  |
|  | 2. Per Month |  |  |  |
|  | 3. Per week |  |  |  |
|  | 5. Per day |  |  |  |
| Fish Intake | 1.rarely/never | 0.059 (0.043, 0.075) | 6.29E-13 | 6.29E-13 |
|  | 2. Per Year |  |  |  |
|  | 2. Per Month |  |  |  |
|  | 3. Per week |  |  |  |
|  | 5. Per day |  |  |  |
| French fries or pan-fried potatoes, poutine | 1.rarely/never | -0.064 (-0.077, -0.053) | 1.01E-24 | 1.01E-24 |
|  | 2. Per Year |  |  |  |
|  | 2. Per Month |  |  |  |
|  | 3. Per week |  |  |  |
|  | 5. Per day |  |  |  |
| Fruit intake | 1.rarely/never | 0.102 (0.078, 0.126) | 2.84E-16 | 2.84E-16 |
|  | 2. Per Year |  |  |  |
|  | 2. Per Month |  |  |  |
|  | 3. Per week |  |  |  |
|  | 5. Per day |  |  |  |
| Green Salad intake | 1.rarely/never | 0.084 (0.065, 0.103) | 2.35E-17 | 2.35E-17 |
|  | 2. Per Year |  |  |  |
|  | 2. Per Month |  |  |  |
|  | 3. Per week |  |  |  |
|  | 5. Per day |  |  |  |
| legumes: beans, peas, lentils intake | 1.rarely/never | 0.094 (0.080, 0.109) | 2.79E-36 | 2.79E-36 |
|  | 2. Per Year |  |  |  |
|  | 2. Per Month |  |  |  |
|  | 3. Per week |  |  |  |
|  | 5. Per day |  |  |  |
| Milk-based dessert intake | 1.rarely/never | -0.011 (-0.022, 0.001) | 0.070311385 | 0.070311385 |
|  | 2. Per Year |  |  |  |
|  | 2. Per Month |  |  |  |
|  | 3. Per week |  |  |  |
|  | 5. Per day |  |  |  |
| Nuts, seeds, and peanut butter intake | 1.rarely/never | 0.079 (0.064, 0.095) | 6.82E-24 | 6.82E-24 |
|  | 2. Per Year |  |  |  |
|  | 2. Per Month |  |  |  |
|  | 3. Per week |  |  |  |
|  | 5. Per day |  |  |  |
| Omega-3 Eggs intake | 1.rarely/never | 0.022 (0.012:0.033) | 6.76E-05 | 6.76E-05 |
|  | 2. Per Year |  |  |  |
|  | 2. Per Month |  |  |  |
|  | 3. Per week |  |  |  |
|  | 5. Per day |  |  |  |
| other vegetables (except carrots, potatoes, or salad) intake | 1.rarely/never | 0.175 (0.152:0.198) | 2.32E-50 | 2.32E-50 |
|  | 2. Per Year |  |  |  |
|  | 2. Per Month |  |  |  |
|  | 3. Per week |  |  |  |
|  | 5. Per day |  |  |  |
| Pastries intake | 1.rarely/never | 0.018 (0.006:0.032) | 0.00406896 | 0.00406896 |
|  | 2. Per Year |  |  |  |
|  | 2. Per Month |  |  |  |
|  | 3. Per week |  |  |  |
|  | 5. Per day |  |  |  |
| pâtés, cretons, terrines | 1.rarely/never | -0.022 (-0.038:-0.007) | 0.005200246 | 0.005200246 |
|  | 2. Per Year |  |  |  |
|  | 2. Per Month |  |  |  |
|  | 3. Per week |  |  |  |
|  | 5. Per day |  |  |  |
| Potatoes intake | 1.rarely/never | -0.041 (-0.059: -0.023) | 2.04E-05 | 2.04E-05 |
|  | 2. Per Year |  |  |  |
|  | 2. Per Month |  |  |  |
|  | 3. Per week |  |  |  |
|  | 5. Per day |  |  |  |
| regular vinaigrettes, dressings, and dips | 1.rarely/never | 0.019(0.006:0.032) | 0.003565544 | 0.003565544 |
|  | 2. Per Year |  |  |  |
|  | 2. Per Month |  |  |  |
|  | 3. Per week |  |  |  |
|  | 5. Per day |  |  |  |
| Salty Snacks intake | 1.rarely/never | -0.002 (-0.015:0.010) | 0.711148033 | 0.711148033 |
|  | 2. Per Year |  |  |  |
|  | 2. Per Month |  |  |  |
|  | 3. Per week |  |  |  |
|  | 5. Per day |  |  |  |
| Sauces and gravies intake | 1.rarely/never | -0.030 (-0.041: -0.019) | 5.96E-07 | 5.96E-07 |
|  | 2. Per Year |  |  |  |
|  | 2. Per Month |  |  |  |
|  | 3. Per week |  |  |  |
|  | 5. Per day |  |  |  |
| Sausages, hot dogs, ham, smoked meat, and bacon | 1.rarely/never | -0.042 (-0.055: -0.030) | 1.61E-10 | 1.61E-10 |
|  | 2. Per Year |  |  |  |
|  | 2. Per Month |  |  |  |
|  | 3. Per week |  |  |  |
|  | 5. Per day |  |  |  |
| Whole Milk intake | 1.rarely/never | -0.013 (-0.031:0.005) | 0.145062784 | 0.145062784 |
|  | 2. Per Year |  |  |  |
|  | 2. Per Month |  |  |  |
|  | 3. Per week |  |  |  |
|  | 5. Per day |  |  |  |
| Low-fat yogurt intake | 1.rarely/never | 0.015 (0.006:0.023) | 0.000763273 | 0.000763273 |
|  | 2. Per Year |  |  |  |
|  | 2. Per Month |  |  |  |
|  | 3. Per week |  |  |  |
|  | 5. Per day |  |  |  |
| Regular Yoghurt intake | 1.rarely/never | 0.016 (0.008:0.026) | 0.000389631 | 0.000389631 |
|  | 2. Per Year |  |  |  |
|  | 2. Per Month |  |  |  |
|  | 3. Per week |  |  |  |
|  | 5. Per day |  |  |  |
| PURE healthy diet score | Total score: 0-28 | 0.024 (0.021: 0.027) | 4.93E-56 | 4.93E-56 |
| PURE healthy diet score Quartiles | Q1: [0-9] | 1 | 4.49E-48 |  |
|  | Q2: (9-13] | 2. 0.128 (0.092:0.165) |  | 4.60E-12 |
|  | Q3: (13-16] | 3. 0.201 (0.161:0.240) |  | 8.11E-23 |
|  | Q4: (16-28] | 4. 0.297 (0.257: 0.338) |  | 1.70E-46 |
| PURE healthy diet score deciles | D1: [0-6] |  | 2.61724E-48 |  |
|  | D2: (6-8] | 0.130 (0.069:0.190) |  | 2.90442E-05 |
|  | D3: (8-10] | 0.167 (0.111:0.223) |  | 7.43234E-09 |
|  | D4: (10-11] | 0.174 (0.108:0.239) |  | 2.30252E-07 |
|  | D5: (11-13] | 0.220 (0.165:0.274) |  | 9.41196E-15 |
|  | D6: (13-14] | 0.260 (0.195:0.325) |  | 9.41196E-15 |
|  | D7: (14-15] | 0.262 (0.196:0.328) |  | 1.59292E-14 |
|  | D8: (15-17] | 0.320 (0.260:0.377) |  | 3.59358E-26 |
|  | D9: (17-19] | 0.380 (0.312:0.441) |  | 8.17811E-30 |
|  | D10: (19-28] | 0.410 (0.344:0.476) |  | 2.90525E-33 |
| Mediterranean diet score | Score (0-50) | 0.0186 (0.0157:0.0215) | 4.90E-36 | 4.90E-36 |
| Mediterranean diet score quartiles | Q1: [3-18] | 1 | 2.07E-25 |  |
|  | Q2: (18-21] | 2. 0.073 (0.035:0.111) |  | 0.000190618 |
|  | Q3: (21-24] | 3. 0.148 (0.109: 0.187) |  | 1.1945E-13 |
|  | Q4: (24-50] | 4. 0.197 (0.159:0.235) |  | 4.99509E-24 |
| Mediterranean diet score deciles | D1: [3-15] | Ref. | 1.38E-28 | Ref. |
|  | D2: (15-17] | 0.129 (0.073:0.184) |  | 6.91427E-06 |
|  | D3: (17-18] | 0.14 (0.074:0.205) |  | 2.97783E-05 |
|  | D4: (18-20] | 0.152 (0.100:0.204) |  | 1.43141E-08 |
|  | D5: (20-21] | 0.15 (0.087:0.212) |  | 4.33003E-06 |
|  | D6: (21-22] | 0.198 (0.136:0.261) |  | 7.67984E-10 |
|  | D7: (22-24] | 0.243 (0.189:0.296) |  | 2.51283E-18 |
|  | D8: (24-25] | 0.245 (0.178:0312) |  | 2.15147E-12 |
|  | D9: (25-27] | 0.248 (0.186:0.310) |  | 7.36103E-15 |
|  | D10: (27-50] | 0.322 (0.263:0.381) |  | 6.90123E-26 |

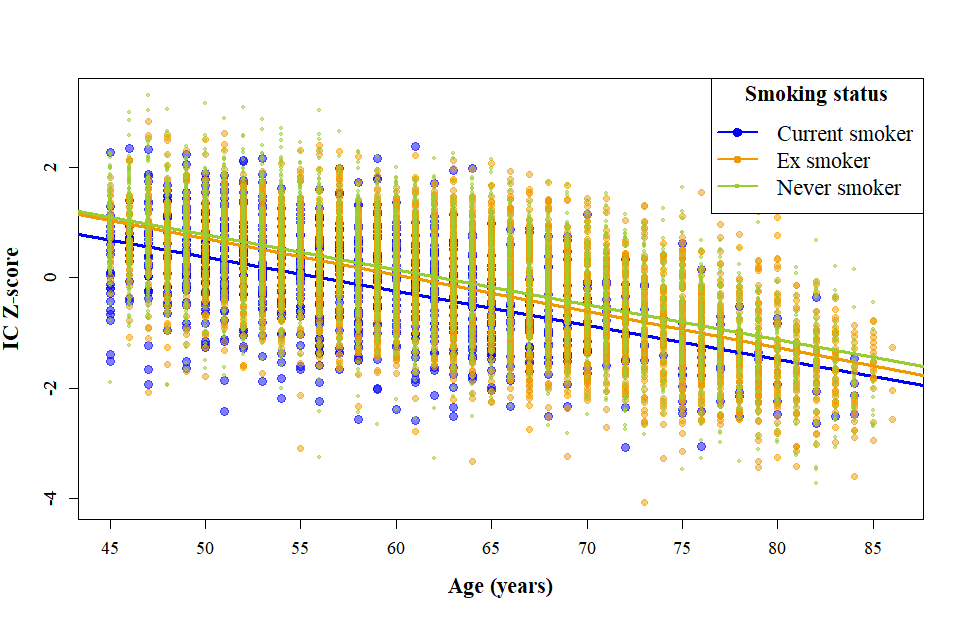

### **Supplementary Figure 1**: Distribution of IC scores across smoking status categories.

*The distribution of IC z-scores (y-axis) across age (x-axis), stratified by smoking status. Separate trend lines are presented for current smokers, ex-smokers, and never smokers. The figure illustrates a general decline in IC scores with increasing age, with lower scores observed among current smokers compared to ex- and never smokers across the age range.*

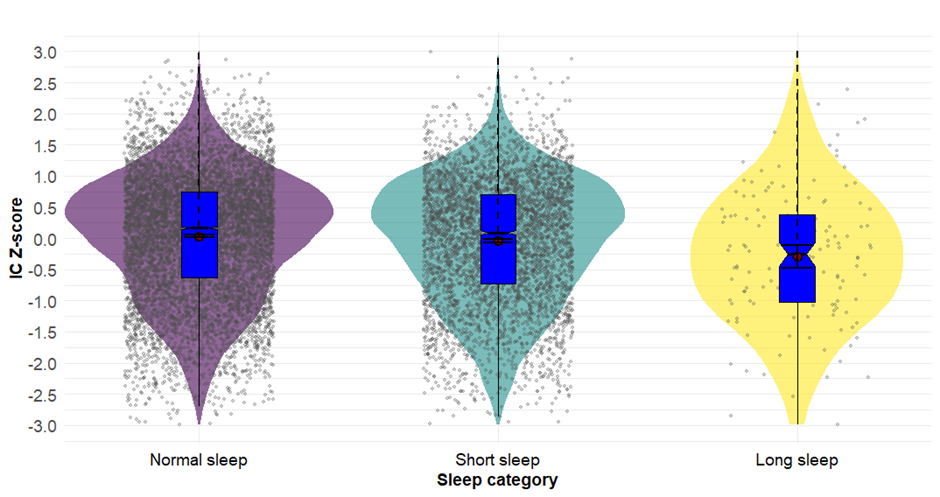

### **Supplementary Figure 2:** Distribution of IC scores by sleep category.

This violin plot illustrates the distribution of IC Z-scores across three sleep duration categories: normal sleep (7–9 hours), short sleep (<7 hours), and long sleep (>9 hours). The y-axis represents the IC Z-score, while the x-axis indicates sleep categories. The width of each violin reflects the density of participants within a given IC score range. Overlaid box plots show the interquartile range and median IC scores for each group. The plot demonstrates lower median IC scores among individuals with short and long sleep durations compared to those with normal sleep duration.

### **Supplementary Table 3**: Socioeconomic and lifestyle factors that showed significant IC associations with PGSxE interaction

|  |  | All Ages | Age < 65 | Age 65+ |
| --- | --- | --- | --- | --- |
| Variable/category | **Categories** | **Beta (95% CI) – PGSxE** | **Beta (95% CI) – PGSxE** | **Beta (95% CI) – PGSxE** |
| Graduated from high school | Graduated vs not | -0.065 (-0.139,0.009) | **-0.109* (-0.211, -0.007)** | -0.021 (-0.153, 0.111) |
| Mediterranean diet score | Score (0-50) | **-0.003* (-0.006, -0.0002)** | -0.003 (-0.006, 0.001) | -0.002 (-0.007, 0.004) |
| Sleep duration | Short vs optimal | -0.021 (-0.049, 0.007) | 0.012 (-0.024, 0.047) | -**0.095* (-0.153, -0.036)** |
|  | Long vs optimal | 0.026 (-0.110, 0.161) | **0.198* (0.023, 0.373)** | -0.191 (-0.459, 0.078) |

### **Supplementary Table 4**: Association of socioeconomic and lifestyle factors stratified by PGS categories (for factors with significant interaction effect)

|  |  | Low PGS | Middle PGS | High PGS |
| --- | --- | --- | --- | --- |
| Variable/category | **Categories** | **Beta (95% CI)** | **Beta (95% CI)** | **Beta (95% CI)** |
| Graduated from high school | Graduated vs not (Age: 45-64) | 0.456 (0.131, 0.781)* | 0.232 (0.117, 0.347)* | 0.024 (-0.255, 0.302) |
|  | Graduated vs not (Age: 65+) | 0.348(-0.055, 0.752) | 0.227(0.107, 0.348)* | 0.408 (0.065, 0.751)* |
| Mediterranean diet score | Score (All Ages) | 0.026(0.017, 0.035)* | 0.018(0.015, 0.022)* | 0.010 (0.002, 0.019)* |
| Sleep duration | Short vs optimal (Age:45-64) | -0.165 (-0.271, -0.060)* | -0.107 (-0.145, -0.071)* | -0.143 (-0.248, -0.039)* |
|  | Short vs optimal (Age: 65+) | -0.088 (-0.252, 0.076) | -0.108 (-0.164, -0.052)* | -0.197 (-0.359, -0.034)* |
|  | Long vs optimal (Age: 45-64) | -0.806 (-1.367, -0.244)* | -0.246 (-0.425, -0.067)* | 0.134 (-0.360, 0.628) |
|  | Long vs optimal (Age: 65+) | 0.32 (-0.576, 0.641) | -0.301 (-0.553, -0.048)* | -0.770 (-1.877, 0.337) |

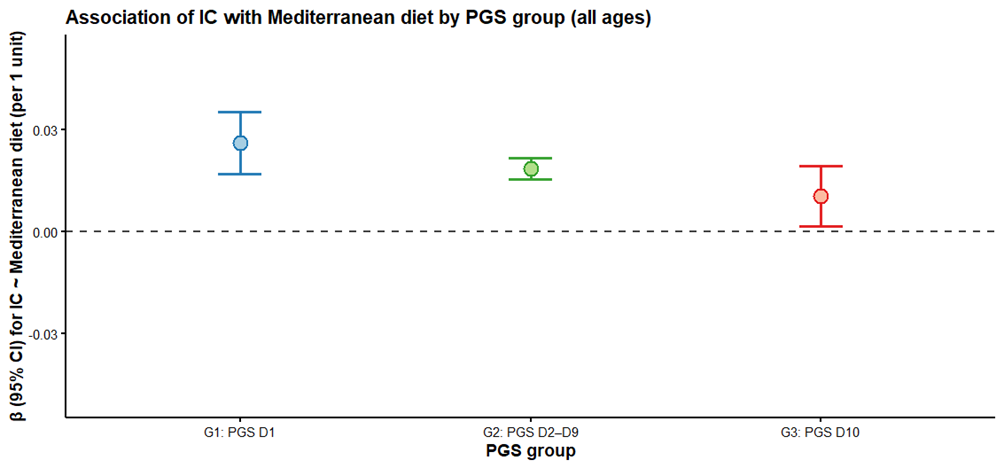

### **Supplementary Figure 3**: Association of IC with Mediterranean diet score across PGS groups (all ages).

Points are linear regression coefficients (β) for IC per 1-unit increase in the Mediterranean diet score, estimated separately within three polygenic score (PGS) groups (G1: D1; G2: D2–D9; G3: D10). Vertical bars are 95% confidence intervals. The dashed horizontal line indicates β = 0.

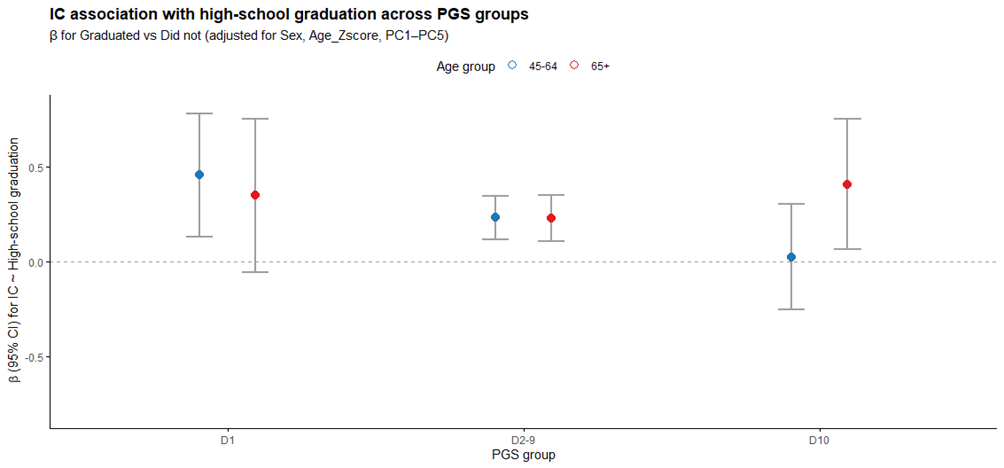

### **Supplementary Figure 4**: Association of IC with high-school graduation by PGS group and age.

Points show linear regression coefficients (β) for IC comparing Graduated vs Did not graduate high school, estimated separately within three polygenic score (PGS) groups (D1, D2–9, D10) and two age strata (45–64 in blue; 65+ in red). Vertical bars are 95% confidence intervals. The dashed horizontal line indicates β = 0.

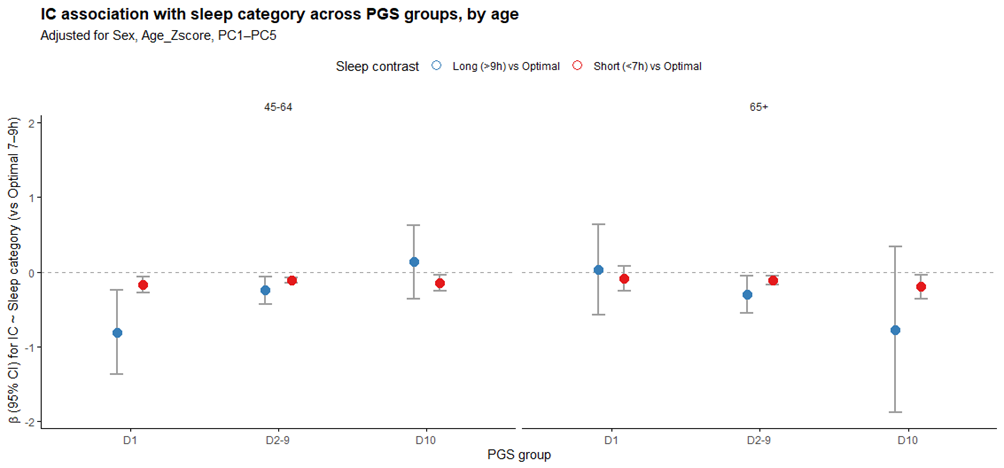

**Supplementary Figure 5**: Association of IC with sleep category across PGS groups, by age.

Points are linear regression coefficients (β) for IC comparing each sleep category with the optimal 7–9 h reference (Short <7 h vs Optimal and Long >9 h vs Optimal). Estimates are shown within three polygenic score (PGS) groups (D1, D2–9, D10) and for two age strata (45–64 and 65+; separate panels). Vertical bars indicate 95% confidence intervals; the dashed horizontal line marks β = 0.

**References**

1.	Mente, A., et al., *Diet, cardiovascular disease, and mortality in 80 countries.* Eur Heart J, 2023. **44**(28): p. 2560-2579.

2.	Bassim, C., et al., *Oral health, diet, and frailty at baseline of the Canadian longitudinal study on aging.* Journal of the American Geriatrics Society, 2020. **68**(5): p. 959-966.

3.	Vahid, F., P. Wilk, and T. Bohn, *Longitudinal effects of diet quality on healthy aging - Focus on cardiometabolic health: findings from the Canadian longitudinal study on aging (CLSA).* Aging Clinical and Experimental Research, 2025. **37**(1): p. 157.

4.	Aoun, C., et al., *Comparison of five international indices of adherence to the Mediterranean diet among healthy adults: similarities and differences.* Nutrition research and practice, 2019. **13**(4): p. 333-343.

5.	Washburn, R.A., et al., *The physical activity scale for the elderly (PASE): Development and evaluation.* Journal of Clinical Epidemiology, 1993. **46**(2): p. 153-162.

6.	Institutes, N.E.R., *PASE: Physical Activity Scale for the Elderly: Administration and Scoring Instruction Manual*. 1991, New England Research Institutes Watertown, MA.

7.	D’Amore, C., et al., *Physical Activity Behaviour in Middle-Aged and Older Canadian Women and Men: An Analysis of the CLSA.* medRxiv, 2025: p. 2025.03.17.25323990.
